# Rapid NGS Analysis on Google Cloud Platform: performance benchmark and user tutorial

**DOI:** 10.1101/2024.12.10.24318826

**Authors:** Eugenio Franzoso, Mariangela Santorsola, Francesco Lescai

## Abstract

Next-Generation Sequencing (NGS) is being increasingly adopted in clinical settings as a tool to increase diagnostic yield in genetically determined pathologies. However, for patients in critical conditions the time-to-results of data analysis is crucial for a rapid diagnosis and response. Sentieon DNASeq and Clara Parabricks Germline are two widely used pipelines for ultra-rapid NGS analysis, but their high computational demands often exceed the resources available in many healthcare facilities. Cloud platforms, like Google Cloud Platform (GCP), offer scalable solutions to address these limitations. Yet, setting up these pipelines in a cloud environment can be complex. This work provides a benchmark of the two solutions, and offers a comprehensive tutorial aimed at easing their implementation on GCP by healthcare bioinformaticians. Additionally, it presents a valuable cost guidance to healthcare managers who consider implementing cloud-based NGS processing. Using five publicly available exome (WES) and five genome (WGS) samples, we benchmarked both pipelines on GCP in terms of runtime, cost, and resource utilisation. Our results show that Sentieon and Parabricks perform comparably. Both pipelines are viable options for rapid, cloud-based NGS analysis, enabling healthcare providers to access advanced genomic tools without the need for extensive local infrastructure.

## Introduction

There are several scenarios where a very rapid diagnosis of a patient can make the difference^1,2^. This is particularly relevant in the case of critically ill paediatric patients who are in intensive care units (PICU and NICU)^3,4^. In these contexts, Next-Generation Sequencing (NGS) has emerged as a crucial tool in the diagnostic process, because it has dramatically increased the diagnostic yield compared to traditional diagnostics^5,6^.

However, while introducing undisputable advantages, NGS increases the burden in data analysis and interpretation leading to diagnosis^7,8^. Therefore, while other steps of the clinical workflow have little margins for speed-ups, innovations in bioinformatics have the potential to dramatically impact the time-to-diagnosis in these scenarios. Amongst the available tools on the market for ultra-rapid NGS data processing are Sentieon^9^ and Clara Parabricks^10^. The first one accelerates the analysis by differently exploiting Central Processing Units (CPUs), while the second one accelerates the workflow by using Graphical Processing Units (GPUs). By accelerating the bioinformatic analysis, these tools allow clinicians to make a faster diagnosis, while also leading to substantial cost savings for healthcare providers^11,12^. Despite these improvements, a major hurdle remains: the requirement of substantial computational resources, such as large multicore servers or GPU cards, often unavailable in many hospital settings. To address this issue, we argue that a cloud-first strategy is a feasible solution in most settings. Cloud computing simplifies the deployment of these advanced software to institutions lacking the necessary infrastructure, while maintaining compliance to regulatory requirements^13^. Besides providing a flexible, maintained and backed-up computing solution, the adoption of a cloud solution makes costs predictable and proportional to the actual demand. For these reasons, we have benchmarked Sentieon and Parabricks to measure their performance in terms of speed and costs on the Google Cloud Platform (GCP). We focused our comparison on key parameters such as sample processing time, cost per sample, CPU and memory usage as well as ease of implementation. The goal of this benchmark is to provide healthcare managers and clinical bioinformaticians with a high-level estimate of resources and a step-by-step guideline to facilitate the implementation of ultra-rapid NGS solutions.

## Materials and Methods

### Samples and Data availability

We assessed the performance of Sentieon and Parabricks for ultra-rapid NGS data processing of human data, using five exome (WXS) and five genome (WGS) samples from the Sequence Read Archive (SRA).

The exome sequencing data we selected belong to a study focused on patients with a syndromic condition characterised by lymphoproliferation, immunodeficiency, and hemophagocytic lymphohistiocytosis (HLH) like phenotypes^14^. The genomic DNA extracted from these patients underwent exome enrichment using the Twist Core Exome capture system. Sequencing was performed on an Illumina NextSeq 500 platform, with a paired-end 75 base pairs (bp) read length.

The genome sequencing data belong to Illumina’s Polaris project, aimed at developing publicly available resources for population genomics analyses^15^. The samples were obtained from sequencing on an Illumina HiSeqX sequencer using a 150bp read length.

Detailed information regarding the individual sample identifiers and corresponding data can be found in **Supplementary Table S1**.

### Design of the benchmark

We processed the ten samples from the raw FASTQ files to VCF using two state-of-the-art, ultra-rapid germline variant calling pipelines: Sentieon’s DNASeq v202308^16^ and CLARA Parabricks Germline v4.0.1-1^17^. To ensure a standardised comparison, both pipelines were launched with their default parameters and execution steps, including alignment, marking duplicates, base recalibration and variant calling.

To accommodate the distinct hardware requirements of each pipeline, we utilised Google Cloud Platform (GCP) to set up two dedicated virtual machines (VMs), each one tailored for either of the pipelines. **Figure 1** illustrates the overall design of our benchmarking analysis, focused on evaluating and comparing the performance of each pipeline in terms of runtime, overall cost, and resource allocation (CPU and memory usage).

**Figure 1:**
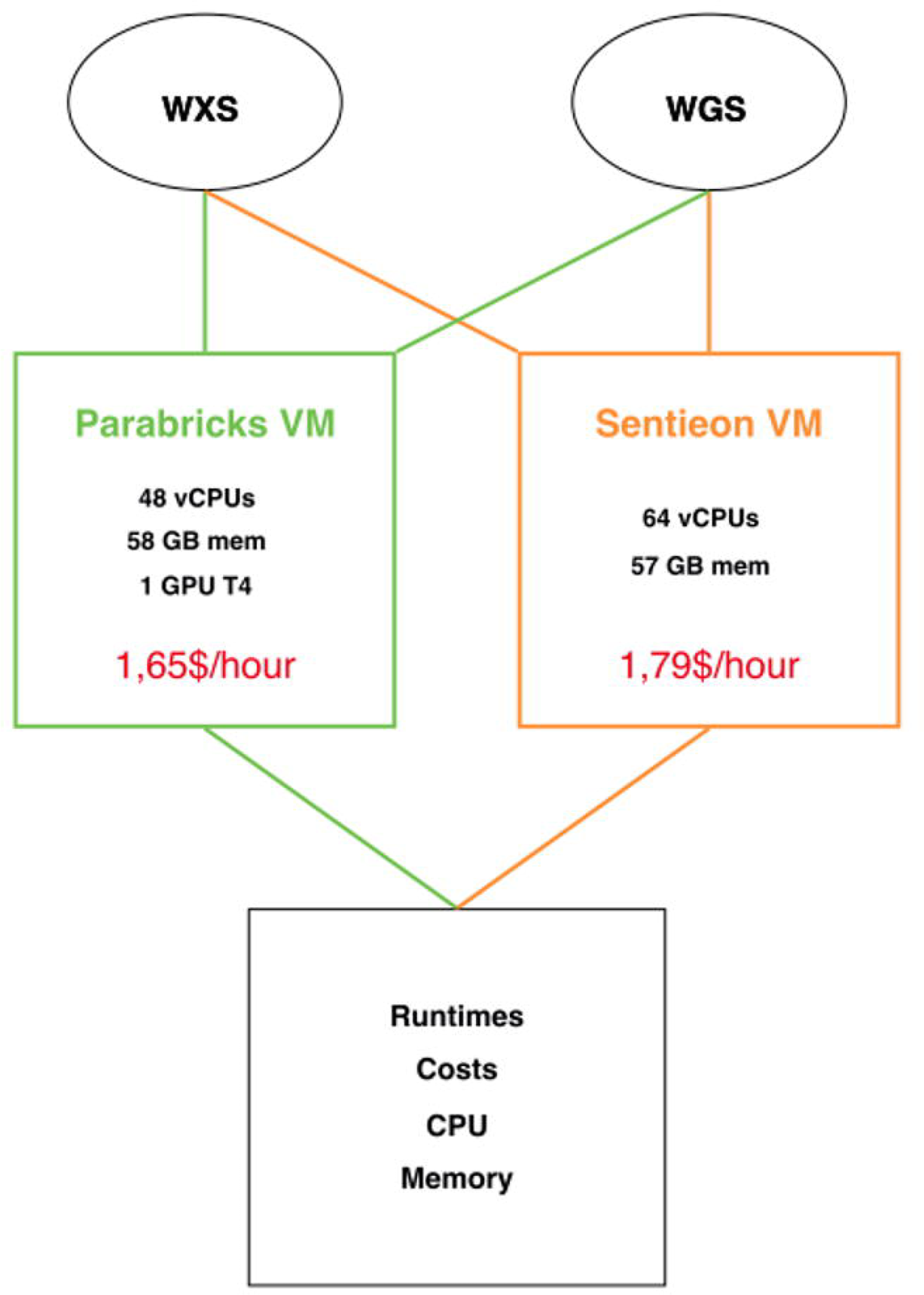
Exome (WXS) and genome (WGS) samples were processed on two distinct VMs by Sentieon and Parabricks, each chosen based on a comparable baseline cost per hour. The benchmarking parameters are indicated in the square at the bottom of the image.

### Cloud configuration

To choose the virtual machines for this benchmark, we aimed at comparable baseline costs per hour, while meeting each software’s requirements. This cost-driven approach was meant to evaluate the performances the two pipelines would achieve in a similar cost- constrained scenario.

The VM for Sentieon was designed with 64 vCPUs and 57GB of memory, aligning with the tool’s CPU-based processing requirements: this VM had a baseline cost of $1.79/hour. The VM for Parabricks was instead configured with 48vCPUs, 58GB of memory, and 1 T4 NVIDIA GPU: this virtual machine had a baseline cost of 1.65$/hour.

Step-by-step instructions to achieve these configurations are given in the **Supplementary file 1**.

### Data collection and analysis

To gather performance metrics, we employed the Ops Agent (https://cloud.google.com/monitoring/agent/ops-agent) on each instance. The GCP monitoring dashboard was used to collect all the metrics, including the computation start and end, CPU and memory usage.

All the plots have been generated using R version 4.3.0 (2023-04-21) inside the RStudio Integrated Development Environment (IDE) (version 2023.06.0+421). The R package tidyverse (version 2.0.0) was used for data manipulation and plotting (https://www.tidyverse.org/).

## Results

### Software Runtimes

The analysis of the WXS runtimes (**Figure 2A**) showed that Parabricks’ Germline completed the variant calling process in approximately 10 to 14 minutes. Sentieon’s DNASeq displayed a similar performance, with analysis duration ranging from 14 to 16 minutes.

**Figure 2:**
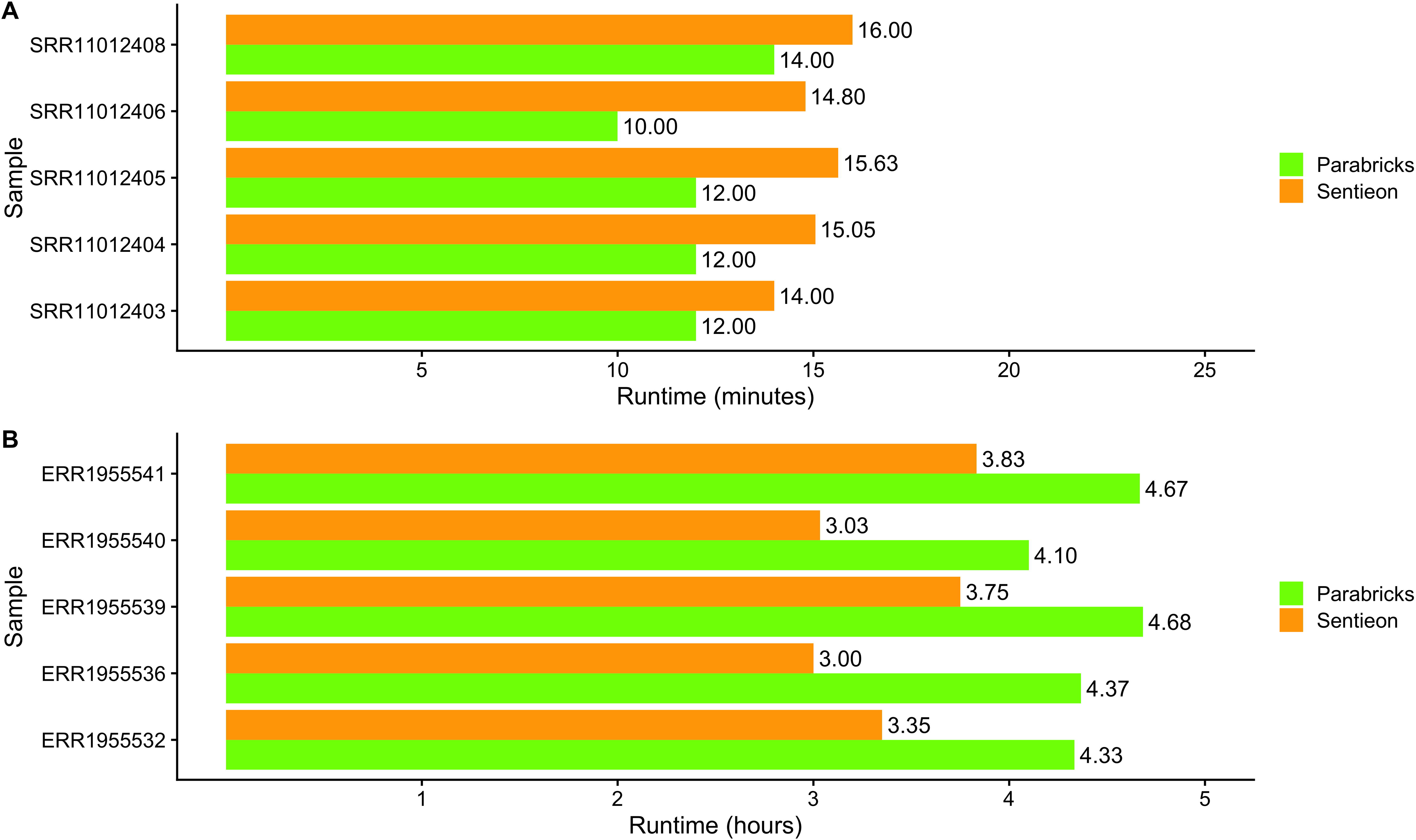
**A)** Runtime analysis (in minutes) of the exome samples (WXS). **B)** Runtime analysis (in hours) of the genome samples (WGS).

As far as the analysis of genome samples, we observed a more significant variation in processing times (**Figure 2B**). The elapsed time for Parabricks analysis ranged between 4.1 and 4.7 hours, while Sentieon analysis lasted between 3 to 3.8 hours.

### Costs

The cost for processing the exome samples with Parabricks Germline ranged between 0.71$ to 0.93$. Using Sentieon the costs of the analysis ranged between 0.82$ and 1.03$ (**Figure 3A**).

**Figure 3:**
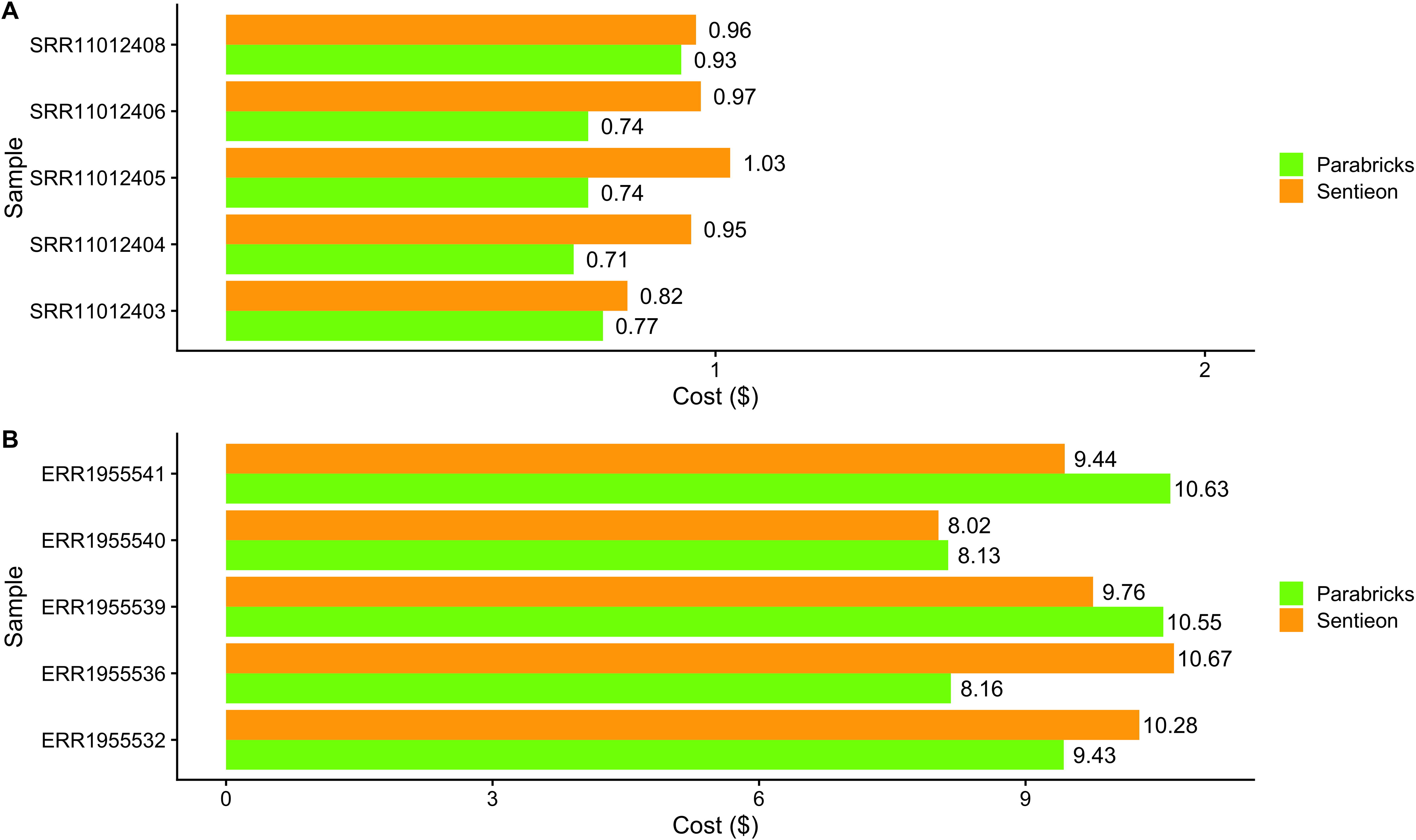
**A)** Cost in USD ($) for the processing of each exome sample. **B)** Cost in USD ($) for the processing of each genome sample.

As far as genome analyses are concerned (**Figure 3B**), the cost of running Parabricks ranged from 8.13$ to 10.63$ per sample while Sentieon’s DNASeq costed between 8.02$ and 10.67$.

Overall, Parabricks Germline incurred lower costs on two out of five WGS datasets (ERR1955532 and ERR1955536).

Given the fundamental differences in the way Sentieon and Parabricks exploit the hardware, and in turn how this impacts these measurements, we also profiled CPU and memory to gather a more complete picture of the overall performance.

### CPU and RAM usage

We profiled in detail both CPU and memory usage throughout the analyses carried out by Sentieon and Parabricks, across all WXS (**Figure 4**) and WGS (**Figure 5**) samples. The analysis of the measurements showed, as expected, that Sentieon is quite greedy in using the available CPUs but not as greedy in using the available memory. On the other hand, Parabricks, despite being accelerated on the GPU which has its own dedicated memory, showed a higher and constant usage of the mainboard memory. At the same time, during the genome samples analysis Parabricks also showed a significant usage of the available (16) CPUs.

**Figure 4:**
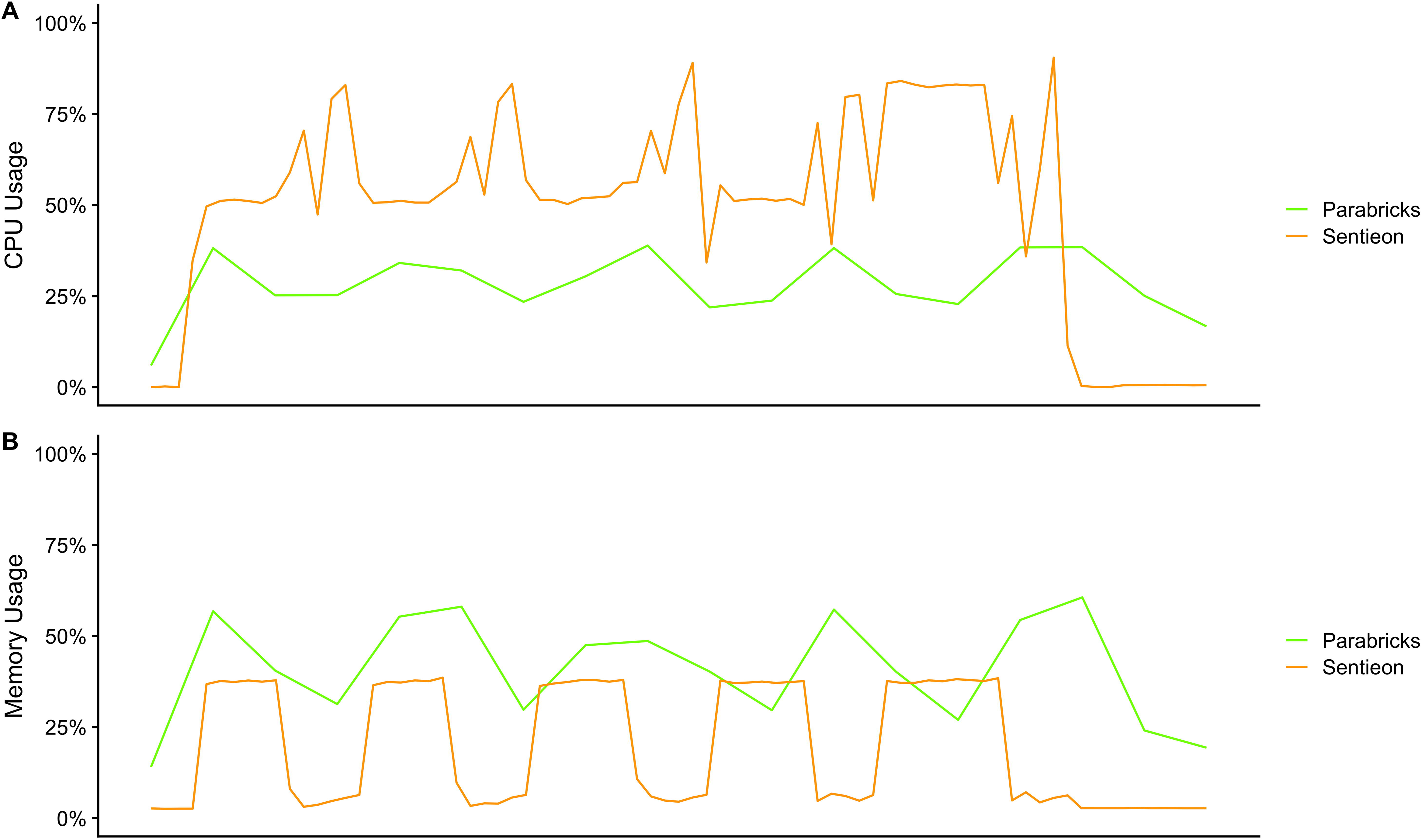
**A)** CPU usage across the five WXS analyses with Sentieon and Parabricks. **B)** Memory usage across the five WXS analyses with Sentieon and Parabricks.

**Figure 5:**
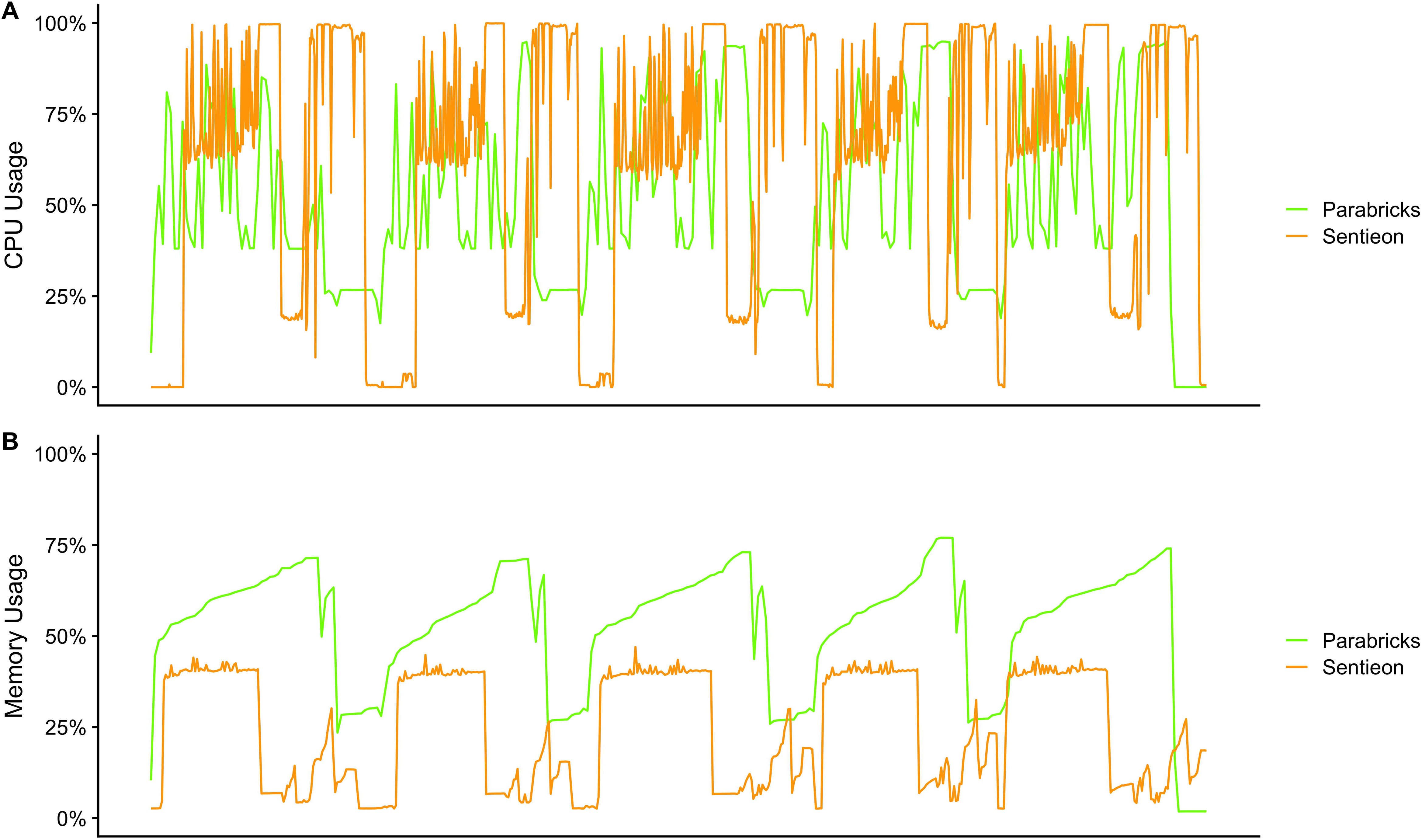
**A)** CPU usage across the five WGS analyses with Sentieon and Parabricks. **B)** Memory usage across the five WGS analyses with Sentieon and Parabricks.

## Discussion

The benchmark presented in this study provides a snapshot of runtime, costs and technical expertise required when performing ultra-rapid NGS data analyses on Google Cloud Platform (GCP).

The decline in the cost of sequencing, along with its high diagnostic yield, has opened new avenues in clinical settings, especially when implemented as a first-line diagnostic test^5,6^. However, to fully leverage these benefits, it is essential to pair sequencing with accurate and rapid bioinformatic approaches, accelerating genetic and physician consultations, and then the route to diagnosis.

Nowadays cloud-based solutions enable healthcare providers to overcome the limitations of on-site hardware, by reducing the time and costs associated with its maintenance and offering scalable and flexible configurations tailored to specific needs^13^. The costs associated with cloud computing are also predictable and directly proportional to the hardware and services chosen, facilitating straightforward budget management.

Our benchmark revealed that, overall, Sentieon and Parabricks on Google Cloud Platform (GCP) yield comparable results in terms of runtime and costs. By standardising the hourly cost of each virtual machine, we ensured a fair comparison of performance metrics across the ten samples analysed in similar a cost-constrained scenario.

While Parabricks performed better than Sentieon in the analysis of exome samples (WXS), Sentieon demonstrated faster runtimes when processing genome ones. This suggests that Sentieon may utilise computational resources more efficiently when handling large datasets in the configurations we tested.

The costs we observed for exome sequencing samples were in line with expectations, with lower prices corresponding to shorter analysis times for both software.

Interestingly, we did not always observe a linear correlation between runtimes and costs when comparing both software (Supplementary Table S2). For instance, samples ERR1955532 and ERR1955536 were less expensive when running Parabricks, despite taking longer than Sentieon to complete.

Based on our results, both Sentieon and Parabricks are viable solutions for performing ultra- rapid NGS on GCP, exhibiting comparable runtimes and costs. However, some important considerations must be addressed when selecting one pipeline over the other.

Firstly, the costs highlighted in our benchmark do not include the licensing fees required for running Sentieon. In contrast, Parabricks has no licensing costs, but it requires a GPU. While deploying a GPU on-premises (HPC) would be expensive, on GCP its cost is included in the hourly price of the virtual machine.

Another important factor is the ease of configuring the virtual machines. Setting up the VM for Sentieon was straightforward, as the software exclusively relies on CPU resources. However, configuring the VM for Parabricks was more complex due to its need for GPU acceleration, which requires appropriate GPU drivers and CUDA (Compute Unified Device Architecture) libraries compatible with the selected GPU: the “Deep Learning on Linux” images on GCP did not work for us, requiring us to configure the entire machine from scratch. The detailed procedures for this custom setup for Parabricks VM and Sentieon VM on GCP are explained in a dedicated section of this paper and require some familiarity with command-line software installation (**Supplementary file**). The CPU usage profiles suggest important recommendations when choosing the machine to be dedicated to each of the software: when running Sentieon, as one might expect, it is important to choose a virtual machine with a sufficient number of cores, which we evaluated to be between 64 and 96; when choosing the virtual machine for Parabricks, one should obviously choose a performant GPU, but also consider choosing a sufficient amount of memory (48-64) and a minimum of 16 CPUs. The recommended number of CPUs for Parabricks seems to be an important requirement: we observed (data not shown) that providing a lower number of CPUs, when analysing genome samples, would drastically slow down the analysis runtimes.

## Conclusion

This benchmark provides a valuable resource for healthcare managers and clinical bioinformaticians seeking to implement ultra-rapid NGS solutions in their institutions, overcoming the limitations of on-site computational resources.

By providing a high-level estimate of the runtimes, costs, CPU, and memory usage for Sentieon and Parabricks on GCP, alongside a step-by-step guideline for custom VM implementation in cost-sensitive scenarios, we aimed to facilitate the widespread adoption of these solutions in clinical settings.

With the adoption of these tools, clinicians will be able to make faster diagnoses and, ultimately, improve patient care.

## Supporting information

supplementary material - user tutorial

## Data Availability

All data produced in the present study are available upon reasonable request to the authors

## Authors’ contributions

Conceptualization: F.L., M.S. and E.F.; Formal analysis: E.F.; Writing-original draft: F.L., E.F. and M.S.; Writing-review & editing: F.L., E.F. and M.S.

## Conflicts of interest

All the authors declare no conflicts of interest.

## Acknowledgements

We acknowledge Sentieon for providing an evaluation licence to carry out performance testing across multiple environments. We acknowledge the Grant CN00000013 “National Centre for HPC, Big Data and Quantum Computing”, funded through “Decreto Direttoriale di concessione del finanziamento” n.1031 del 17.06.2022 on Next Generation EU funding (PNRR MUR) – M4C2 – Investimento 1.4 - Avviso “Centri Nazionali” -D.D. n. 3138 dated 16^th^ December 2021

## Supplementary Materials

**Supplementary Table S1**: Size and identifiers of the publicly available samples used for the benchmark.

**Supplementary Table S2**: Runtime and costs for each of the samples analysed.

**Supplementary Materials SM1**: User Tutorial. Step-by-step tutorial on how to deploy either Sentieon rapid NGS or NVIDIA Clara Parabricks on Google Cloud Platform.

## Notes

### Competing Interest Statement

The authors have declared no competing interest.

