## supplementary material - user tutorial for "Rapid NGS Analysis on Google Cloud Platform: performance benchmark and user tutorial"

### Supplementary Material SM1: User Tutorial

#### Deployment instructions

In this section we provide a step-by-step configuration guideline to enable experienced and inexperienced users alike to run any of the two software on the Google Cloud Platform. To follow this guide, we assume the reader will have:

a) An account on GCP and a project connected to a valid billing account
b) A minimal experience with the bash shell
c) A valid licence for running Sentieon

**Setup Sentieon analysis**

1. Once the licence has been purchased, you will receive the links for downloading it together with the software. Use the links you have received to download and save both the licence file and the software folder on your machine. These will be transferred later to the VM using a Secure Copy Protocol (SCP).
2. Go to console.cloud.google.com/compute/instances. Click on the "CREATE INSTANCE" button. You will be redirected to a new page where you can choose how to configure your instance.
3. Name the virtual machine and select its region and zone depending on your location. In our case we respectively selected europe-west4 and europe-west4-a as region and zone. Click on the "MANAGE TAGS AND LABELS" dropdown and add a label to the machine. We used "machine" as key and "sentieon" as value.
4. In the "Machine Configuration" section, select the N1 series. Under "Machine Type", select the preset n1-highcpu-64 (64vCPU, 32 core, 57.6 GB memory).
5. Under the Boot Disk section, click on "CHANGE". Select CentOS public image, version CentOS 7. Leave the boot disk type untouched and choose the size (GB) of the hard disk depending on your preference. We opted for 500 GB.
6. Check the "Install Ops Agent for monitoring and logging" under the Observability – Ops Agent section.
7. Now you can finally click on "CREATE" at the bottom of the page and wait for the VM instantiation.

8. Go back to console.cloud.google.com/compute/instances, and you should see the created virtual machine. This may take a while. Click on SSH to connect to the virtual machine. A new pop-up window will open with a shell on the machine.
9. Now you will be able to install the required software, using the following command:
   `sudo yum install git screen bzip2 libxml2-dev wget`
10. To transfer files from a local infrastructure to GCP, a ssh key must be added to `~/.ssh/authorized_keys` on the VM. On your local machine, create a public key for transferring Sention's licence and software folder to the VM. Open your terminal window and type:
    `ssh-keygen -t rsa -f ~/.ssh/gcloud_key -C /local_username -b 2048`
    This will create a public key in your local `~/.ssh` folder, named `gcloud_key.pub`. Use the `cat` command to visualise the key and copy it entirely:
    `cat ~/.ssh/gcloud_key.pub`
11. Go back to the VM command line and edit the `~/.ssh/authorized_keys` file, e.g.:
    `vim ~/.ssh/authorized_keys`
12. Paste the key you have just copied on your local machine in the file, making sure that the format is the same as the other keys that have been added automatically during the instantiation of the VM by Google. You will have to modify the last part of the key, where your username is specified. To save and exit the file once modified, type `:wq`. This is an example of the format required:
    
    ```
    ssh-rsa
    AAeAB3NzaC1yc2EAAAADAQABAAABAQC7NX4iKXDXYsfFEac9QSQSI6xz5H69etM+G
    IKhQjx5MhbyZcaR/fWPxhU6DK9vu8PIaZOMN7ilh6k/mllC1LZnLnibWw5z8Vrw1S
    PPaDF56SH1hcWa1+1pgPG+89v1jzKl8rJLqfOlXkISSYGjWXY/yvo6N/M2pDq/XU7
    uh3txFhGcam9BcUxnsqJhnPCsHv/+P9A1eowOIkcFazJ+y3ctg9MhhC/jblGVQLEw
    l6nCFsZpsVZyFwaB8kKV95wnD9SvGHHPYWphIJPASKtr+qFSGt75H4yrjRgkCRYH0
    2JQgpLfW1jo/sPs0c4iKaVhO1oi8uaqJL8tTFJtKGOHvFHz local_username
    {"userName":"local_username","expireOn":"YYYY-MM-
    00T09:58:35+0000"}
    ```
    
13. Go back to your home with `cd ~` and create two new folders: one for the Human genome references and one for the sample.
    `mkdir -p references`
    `mkdir -p sample`

14. Now you can copy the required files on the VM from your local server using the SCP protocol. This code snippet shows how to transfer the reference files. Back to your terminal:

```
scp -r -i /home/user/.ssh/gcloud_key
/home/path/to/reference/files
gcloud_username@external_ip_address:/home/gcloud_username/referen
ces/.
```

The same command must be executed to transfer the sample, software licence and software folder to the VM. We copied the software folder and the licence file inside the `/home` folder on the VM, while the sample was copied inside the `/sample` folder.

15. Go to the VM terminal and unzip the software folder:

```
tar xvzf sentieon-genomics-202308.02.tar.gz
```

16. Now you need to move the software licence in a directory named `LICENCE_DIR`.

Type the following commands:

```
mv LICENCE_FILE.lic LICENSE_DIR/.
export SENTIEON_LICENSE=LICENSE_DIR/LICENSE_FILE.lic
```

17. Sentieon provides sample scripts to run exome (wes-interval.sh) or genome (wgs.sh) workflows. You can find them at https://github.com/Sentieon/sentieon-scripts/blob/master/example_pipelines/germline/DNAseq.

Our customised scripts are available at https://github.com/lescailab/genomics-benchmarks/tree/main/sentieon/sentieon_gcp/scripts.

18. Download the script you need, edit it and transfer it to the VM with the following command:

```
scp -r -i /home/user/.ssh/gcloud_key /home/path/to/script/file
gcloud_username@external_ip_address:/home/gcloud_username/.
```

19. Once you have the script on the VM, open a new screen window by typing the command `screen` in your VM terminal. Screen is a terminal multiplexer that allows you to open virtual terminals inside of your session. This means that even if your connection with the VM was interrupted, the processes running in Screen would continue to run. If you want to detach from the screen session, just press the keys `ctrl+a+d`.

20. Once Screen is started, you can launch the software with:

```
bash <wes-interval.sh><wgs.sh>
```

**Parabricks**

1. Go to console.cloud.google.com/compute/instances. Click on the "CREATE INSTANCE" button. You will be redirected to a new page where you can choose how to configure your instance.
2. Name the virtual machine and select its region and zone depending on your location. In our case we respectively selected europe-west4 and europe-west4-a as region and zone. Click on the "MANAGE TAGS AND LABELS" dropdown and add a label to the machine. We used "machine" as key and "parabricks" as value.
3. In the "Machine Configuration" section, select the GPUs series. Choose 1 NVIDIA T4. Under "Machine Type", click on the "CUSTOM" button. Select 48 vCPU cores and 58 GB of memory.
4. Under the Boot Disk section, click on "CHANGE". Select Debian public image, version Debian GNU/Linux 10 (buster). Leave the boot disk type untouched and choose the size (GB) of the hard disk depending on your preference. We opted for 600 GB.
5. **DO NOT** check the "Install Ops Agent for monitoring and logging" under the Observability – Ops Agent section. We have experienced that this might cause an error once the VM is up and running. It will be installed later.
6. Click on the "CREATE" button at the bottom of the page and wait for the VM instantiation.
7. Go back to console.cloud.google.com/compute/instances, and you should see the created virtual machine. This may take a while. Click on SSH to spin it up and open a connection with the machine. You should see a new window from which you can interact with the VM.
8. Install the Ops Agent. From your `/home` folder:
   ```
   curl -sSO https://dl.google.com/cloudagents/add-google-cloud-ops-agent-repo.sh
   sudo bash add-google-cloud-ops-agent-repo.sh --also-install
   ```
9. To use the GPU, the correct drivers must be installed on the system. If you have selected an NVIDIA T4 GPU as indicated in step 3, then just type the following commands in your VM:
   ```
   # Ensure Python3 is installed on the system
   python3 --version
   ```

```
# Download the startup script
curl
https://raw.githubusercontent.com/GoogleCloudPlatform/compute-gpu-installation/main/linux/startup_script.sh --output
startup_script.sh

# Download the installation script
curl
https://raw.githubusercontent.com/GoogleCloudPlatform/compute-gpu-installation/main/linux/install_gpu_driver.py --output
install_gpu_driver.py

# Launch the startup script
sudo bash startup_script.sh

# Check for drivers' installation
sudo nvidia-smi
```

10. Since Parabricks can be downloaded as a container from the web, you need to install Docker on the VM. Type the following commands:

```
# Delete the outdated packages
sudo apt-get purge docker lxc-docker docker-engine docker.io

# Update the default repository
sudo apt-get update

# Download the following dependencies
sudo apt-get install apt-transport-https ca-certificates curl
gnupg2 software-properties-common

#  Download Docker’s official GPG key to verify the integrity of
packages before installing
curl -fsSL https://download.docker.com/linux/debian/gpg | sudo
apt-key add -

# Add the Docker repository to your system repository
sudo add-apt-repository "deb [arch=amd64]
https://download.docker.com/linux/debian buster stable"

# Update the apt repository
sudo apt-get update

# Install Docker Engine – Community (the latest version of
Docker) and containerd
sudo apt-get install docker-ce docker-ce-cli containerd.io

# The service will start automatically after the installation.
Check the status
```

```
sudo systemctl status docker

# Check Docker version
docker --version
```

11. Download the Parabricks container using Docker:

```
docker pull nvcr.io/nvidia/clara/clara-parabricks:4.0.0-1
```

12. Install the NVIDIA Container Toolkit:

```
# Configure the repository
curl -fsSL https://nvidia.github.io/libnvidia-container/gpgkey |
sudo gpg --dearmor -o /usr/share/keyrings/nvidia-container-
toolkit-keyring.gpg \
  && curl -s -L https://nvidia.github.io/libnvidia-
container/stable/deb/nvidia-container-toolkit.list | \
sed 's#deb https://#deb [signed-by=/usr/share/keyrings/nvidia-
container-toolkit-keyring.gpg] https://#g' | \
sudo tee /etc/apt/sources.list.d/nvidia-container-toolkit.list \
  && \
   csudo apt-get update


# Install the NVIDIA Container Toolkit packages
sudo apt-get install -y nvidia-container-toolkit
```

13. Now you need to install the Screen software.  To install it:

```
sudo apt-get install screen
```

14. You can follow steps 10-14 from the Sentieon instructions here, because the process for Parabricks is almost identical. **Important! In the case of Parabricks there is no need for a licence and you are going to use the container downloaded in step 11 to run the software. You only need your sample and reference files on the VM.**
15. Once you have the reference files and sample on your VM, the software can be launched inside a new screen with:

```
sudo docker run \
  --gpus all \
  --rm \
  --volume $(pwd):/workdir \
```

```
  --volume $(pwd):/outputdir \
nvcr.io/nvidia/clara/clara-parabricks:4.0.0-1 \
pbrun germline \
  --ref /workdir/references/Homo_sapiens_assembly38.fasta \
  --in-fq /workdir/sample/$(basename "$fwd")
/workdir/sample/$(basename "$rev") \
  --knownSites
/workdir/references/Homo_sapiens_assembly38.known_indels.vcf.gz \
  --out-bam /outputdir/"${sample_name}"_markdup.bam \
  --out-variants /outputdir/"${sample_name}".vcf \
  --out-recal-file /outputdir/recal.txt \
  --tmp-dir /outputdir/tmp
```

The script we have used is here: https://github.com/lescailab/genomics-benchmarks/blob/main/parabricks/parabricks_gcp/no_loop_runs/launch_parabricks.sh.
